# Rate of Antibiotic Use and Associated Risk Factors in COVID-19 Hospitalized Patients

**DOI:** 10.1101/2020.10.21.20217117

**Authors:** Alysa J. Martin, Stephanie Shulder, David Dobrzynski, Katelyn Quartuccio, Kelly E. Pillinger

**Author notes:** **Conflict of Interest Statement:** The authors have no personal conflicts of interests to disclose. The authors would like to disclose that K. Quartuccio’s spouse consults for AstraZeneca.

## Abstract

**Background:** Literature suggests that antibiotic prescribing in COVID-19 patients is high, despite low rates of confirmed bacterial infection. There are little data on what drives prescribing habits.

**Objective:** This study sought to determine antibiotic prescribing rates and risk factors for antibiotic prescribing in hospitalized patients. It was the first study to assess risk factors for receiving more than one course of antibiotics.

**Methods:** This was a retrospective, multi-center, observational study. Patients admitted from March 1, 2020 to May 31, 2020 and treated for PCR-confirmed COVID-19 were included. The primary endpoint was the rate of antibiotic use during hospitalization. Secondary endpoints included risk factors associated with antibiotic use, risk factors associated with receiving more than one antibiotic course, and rate of microbiologically confirmed infections.

**Results:** A total of 208 encounters (198 patients) were included in the final analysis. Eighty-three percent of patients received at least one course of antibiotics, despite low rates of microbiologically confirmed infection (12%). Almost one-third of patients (30%) received more than one course of antibiotics. Risk factors identified for both antibiotic prescribing and receiving more than one course of antibiotics were more serious illness, increased hospital length of stay, intensive care unit admission, mechanical ventilation, and acute respiratory distress syndrome.

**Conclusion and relevance:** There were high rates of antibiotic prescribing with low rates of bacterial co-infection. Many patients received more than one course of antibiotics during hospitalization. This study highlights the need for increased antibiotic stewardship practices in COVID-19 patients.

## Introduction

The novel coronavirus disease identified in 2019 (COVID-19) is caused by a betacoronavirus known as SARS-CoV-2. Despite the fact that COVID-19 is a viral infection, clinical manifestations may present similarly to a bacterial pneumonia.^1^ Patients often present with respiratory symptoms including fever, cough, dyspnea, and bilateral changes on chest imaging.^1^ It can be difficult to determine if patients have a bacterial respiratory coinfection along with COVID-19 infection. This may lead to over-prescribing of antibiotics in patients with COVID-19.

Despite bacterial coinfection being reported in 2-8% of patients, rates of antibiotic prescribing have ranged from 57-95% in hospitalized patients.^1-7^ Additionally, there are reports of increased consumption of antibiotics during the COVID-19 pandemic.^6-9^ Over-prescribing of antibiotics may increase the risk of adverse side effects, nosocomial infection, and antibiotic resistance. Several organizational guidelines recommend starting empiric antibiotic therapy if concern for bacterial pneumonia or sepsis, but to re-evaluate daily and de-escalate or discontinue if no evidence of bacterial infection.^10-14^ Furthermore, they emphasize the importance of antimicrobial stewardship programs which may help to avoid starting antibiotics reflexively or continuing unnecessary courses. The World Health Organization (WHO) has released guidance which discourages the use of antibiotic therapy for patients with COVID-19 infections unless signs and symptoms of a bacterial infection exist.^11^

Unfortunately, there are limited data to date describing the need or role for antibiotics in COVID-19 or outcomes for patients who receive them. Additionally, little is known regarding risk factors associated with antibiotic use. There have only been two previous studies which evaluated prescribing trends and risk factors of antibiotic prescribing. Both studies focused on prescribing rates, coinfections, and risk factors associated with antibiotic prescribing within the first 48 hours of admission.^4, 5^

The purpose of this study was to evaluate the rate of antibiotic use in COVID-19 patients and risk factors associated with antibiotic use throughout hospitalization. A better understanding of prescribing practices can be used to evaluate opportunities for antibiotic stewardship for future patients with COVID-19.

## Methods

### Study design

This was a multi-center, retrospective, observational study of antibiotic prescribing trends in patients with confirmed COVID-19 at three hospitals within the University of Rochester Medical Center.

Patients 18 years and older admitted from March 1, 2020 to May 31, 2020 and treated for PCR-confirmed COVID-19 were eligible for inclusion. Patients did not meet inclusion criteria if they were only treated in the emergency department without subsequent admission. Patients could be included more than once if they had multiple admissions within the study period. Patients were excluded if they were asymptomatic. Data were collected until June 30, 2020. This study was reviewed and deemed exempt by the Institutional Review Board.

### Data Collection

Patients were identified from the electronic health record based on a reported positive SARS-CoV-2 PCR test. Baseline data including age, gender, race, weight, height, comorbidities, date of first positive SARS-CoV-2 PCR, and admission and discharge dates were collected. Labs on admission (or at time of positive test if nosocomial transmission), microbiologic data, clinical symptoms, imaging, antibiotic data, COVID-19 investigational or off-label treatment, death and re-admission within 30 days were also collected.

### Outcomes

The primary endpoint of the study was the rate of antibiotic use in patients admitted for COVID-19. Secondary endpoints included risk factors associated with antibiotic use, risk factors associated with receiving more than one antibiotic course, and rate of microbiologically-confirmed infections. Microbiologically-confirmed infection was defined as a positive result from positive urinary antigens or respiratory and blood cultures not suggestive of contaminant or colonization.

### Statistical Analysis

Data were analyzed using R statistical software version 4.0.2 (Boston, MA) with a p-value < 0.05 considered statistically significant. Continuous variables were described using a median with interquartile range (IQR). Categorical variables were analyzed using Fisher’s exact testing. Two-sample Wilcoxon test was used for continuous variables. To evaluate risk factors associated with antibiotic use, a univariate analysis was conducted and variables with a p-value ≤ 0.05 were included in a multivariate logistic regression.

## Results

A total of 228 encounters met inclusion criteria, with twenty excluded due to asymptomatic disease, leaving 208 encounters included in the final analysis (n=198 patients). The median age was 69 years (IQR 60-80) and 50% (105/208) were male. The most common comorbidities were hypertension (143/208, 69%), obesity (81/208, 39%), and diabetes (81/208, 39%). Of the 208 encounters, 83% (172/208) of patients received at least one course of antibiotics (Table 1). Antibiotic prescribing did not differ based on gender, age, race, or comorbidities. Even though a small subset of patients (n=11), all immunocompromised patients received antibiotics

**Table 1.** Baseline Characteristics of COVID-19 patients and Risk factors for Antibiotic Prescribing.

| **Baseline Characteristic** | **Total** | **Antibiotics**<br>**n=172** | **No Antibiotics**<br>**n=36** | **P-value** |
| --- | --- | --- | --- | --- |
| Male gender | 105 (50.5) | 84 (48.8) | 21 (55.6) | 0.43 |
| Age, years (median, IQR) | 69.0 (60-80) | 70.0 (61-81) | 65.5 (47-77) | 0.12 |
| Race |  |  |  | 0.56 |
| Caucasian | 116 (55.7) | 98 (56.9) | 18 (47.2) |  |
| Black/African American | 69 (33.2) | 53 (30.8) | 16 (44.4) |  |
| Asian | 2 (1.0) | 2 (1.2) | 0 (0) |  |
| Other | 6 (2.9) | 6 (3.5) | 0 (0) |  |
| Unknown | 15 (7.2) | 13 (7.6) | 2 (5.6) |  |
| Comorbidities |  |  |  |  |
| Hypertension | 143 (68.8) | 116 (67.4) | 27 (72.2) | 0.43 |
| Diabetes | 81 (38.9) | 70 (23.2) | 11 (27.8) | 0.35 |
| Obesity | 80 (38.5) | 66 (38.4) | 14 (38.9) | >0.99 |
| Cardiac Disease | 56 (26.9) | 50 (29.1) | 6 (16.7) | 0.15 |
| Chronic Lung Disease | 43 (20.7) | 37 (21.5) | 6 (16.7) | 0.65 |
| CKD | 45 (21.7) | 40 (23.2) | 5 (13.9) | 0.20 |
| None | 21 (10.1) | 17 (9.9) | 4 (11.1) | 0.77 |
| Immunocompromised<sup>a</sup> | 11 (5.3) | 11 (6.4) | 0 (0) | 0.22 |
| Labs (median, IQR) |  |  |  |  |
| AST, U/L | 43 (31-65) | 46 (33.25-65) | 35 (26.75-49) | 0.02 |
| ALT, U/L | 30 (20-46) | 29.5 (20-45) | 32.5 (20-49.5) | 0.83 |
| Scr, mg/dL | 1.130 (0.84-1.64) | 1.14 (0.84-1.725) | 1.06 (0.81-1.3) | 0.14 |
| WBC, /mm<sup>3</sup> | 7.30 (5.50-10.50) | 7.4 (5.5-10.7) | 6.8 (4.8-9.2) | 0.11 |
| ALC, K/uL | 1.0 (0.7-1.3) | 1.0 (0.7-1.3) | 1.1 (0.5-1.4) | 0.10 |
| CRP, mg/L | 93.0 (39.5-134.5) | 93 (39.5-134.5) | 73 (37.5-118.5) | 0.37 |
| LDH, U/L | 344 (264.5-467) | 348 (285-474) | 294 (239-387) | 0.03 |
| Ferritin, ng/mL | 627 (304-1086) | 627 (309-1133.5) | 589 (235-980) | 0.45 |
| D-dimer, ug/mL | 1.26 (0.63-2.08) | 1.37 (0.71-2.1) | 0.86 (0.6-1.6) | 0.24 |
| PCT, ng/mL | 0.215 (0.120-0.620) | 0.28 (0.13-0.63) | 0.13 (0.09-0.20) | <0.01 |
| Symptoms Prior to Admission |  |  |  |  |
| Fever | 120 (57.7) | 101 (58.7) | 19 (52.8) | 0.59 |
| Respiratory Symptoms | 180 (86.5) | 152 (88.4) | 28 (75) | 0.10 |
| Gastrointestinal symptoms | 76 (36.5) | 60 (34.9) | 16 (41.7) | 0.34 |
| Anosmia/Hyposmia/Hypogeusia | 11 (5.3) | 10 (5.8) | 1 (2.8) | 0.69 |
| Symptom duration prior to admission, days (median, IQR) | 6.0 (3-9) | 6.0 (3-9) | 7.5 (4.75-9) | 0.16 |
| Location prior to admit |  |  |  | 0.08 |
| Home | 140 (67.3) | 110 (63.9) | 30 (80.5) |  |
| SNF | 60 (28.8) | 54 (31.4) | 6 (16.7) |  |
| Transfer from OSH | 8 (3.8) | 8 (4.7) | 0 (0) |  |
| Duration of admission, days (median, IQR) | 11 (5-22) | 12 (5.75-25) | 5.5 (4-11.3) | <0.01 |
| ICU admission | 104 | 102 (58.9) | 2 (5.6) | <0.01 |
| ICU LOS, days (median, IQR) | 10 (3-18) | 10 (3.25-18) | 6.5 (6.25-6.75) | 0.59 |
| O2 support |  |  |  | <0.01 |
| Room Air | 40 (19.2) | 24 (13.9) | 16 (44.4) |  |
| Standard Nasal Cannula | 67 (32.2) | 49 (28.5) | 18 (50.0) |  |
| HFNC/Venti Mask | 19 (9.1) | 17 (9.9) | 2 (5.6) |  |
| BiPAP | 5 (2.4) | 5 (2.9) | 0 (0) |  |
| Non-rebreather | 7 (3.4) | 7 (4.1) | 0 (0) |  |
| Mechanical ventilation | 70 (33.7) | 70 (40.7) | 0 (0) |  |
| ARDS | 78 (37.5) | 78 (45.3) | 0 (0) | <0.01 |
| Received investigational or off-label treatment for COVID-19<sup>b</sup> | 37 (17.8) | 35 (20.2) | 2 (5.6) | 0.03 |
| 28-day mortality | 51 (24.5) | 48 (27.9) | 3 (8.3) | 0.02 |
| 30-day readmission | 16 (7.7) | 12 (6.9) | 4 (11.1) | 0.75 |
| More than 1 admission for COVID-19 | 12 (5.8) | 8 (4.7) | 4 (11.1) | 0.23 |
| Time from discharge to COVID-19 readmission, days (median, IQR) | 7 (4.5-12.25) | 5 (2.75-11.5) | 10 (7.75-12.5) | 0.22 |

(%) = Percentages unless otherwise specified
a = immunocompromised patients were defined as those with previous solid organ transplant, active cancer receiving chemotherapy, patients on biologic therapy, and uncontrolled HIV
b = Investigational or off-label therapy included remdesivir, remdesivir + baricitinib, hydroxychloroquine, tocilizumab or convalescent plasma
IQR = Interquartile Range
CKD = chronic kidney disease
AST = Aspartate aminotransferase
ALT = Alanine aminotransferase
SCr = Serum Creatinine
WBC = White blood cell count

While rates of antibiotic prescribing overall were high, we saw a temporal decrease in antibiotic prescribing. When consolidating encounters by month of admission, rates of antibiotic prescribing were highest in March, with 88% (42/49) of encounters receiving antibiotics, compared to 82% (127/155) in April and 75% (3/4) in May.

Rates of antibiotic prescribing were significantly higher in patients admitted to the intensive care unit (ICU), requiring O2 support (including ventimask, BiPAP and mechanical ventilation), and with acute respiratory distress syndrome (ARDS) (p < 0.01). Median length of stay was 6.5 days longer in those receiving antibiotics (p < 0.01). Patients who received investigational or off-label therapy for COVID-19 were also more often prescribed antibiotics (p = 0.03). Patients who died within 28 days were more often prescribed antibiotics (92%, 47/51, p = 0.02). Procalcitonin (PCT) at baseline was higher in those who were prescribed antibiotics (p < 0.01). In a subgroup analysis, PCT > 0.5 was associated with a higher rate of antibiotic prescribing [94% (44/47) prescribed antibiotics vs. 6% (3/47) did not receive antibiotics, OR 3.76, CI 1.09-20.1, p = 0.03]. Of patients with a PCT > 0.5, 43/47 had respiratory or blood cultures obtained. Microbiologically confirmed bacterial infection was present in 12% of patients with PCT >0.5 (5/43).

Respiratory cultures were obtained in 32% (67/208) of patients. In total, 19% (13/67) were positive (Table 2). *Staphylococcus aureus* was identified in 5 of 13 positive respiratory culture results (3% of total patients). Blood cultures were obtained in the majority of patients (149/208, 72%), but few were positive (12/149, 7%). All influenza PCR and *Legionella* urinary antigen (UAg) tests were negative (n= 88 and 93 respectively). Two *Streptococcus pneumoniae* UAg tests were positive (2.4%, 2/84). In addition, 75 patients (36%) were screened for MRSA nares colonization. Overall, there were 24 microbiologically-confirmed infections and all were treated with antibiotics [12% (24/208)]. In a subgroup analysis, patients with microbiologically-confirmed infection were more likely to receive additional courses of antibiotics (20/24, 83% p < 0.01), be admitted to the ICU (23/24, 96%, p < 0.01), and have a longer length of stay (median 29.5 days, p < 0.01).

**Table 2:** Microbiologic Data.

| **Table 2: Microbiologic Data** |  |  |  |
| --- | --- | --- | --- |
| **Micro Data** | **Antibiotics**<br>**n=172** | **No antibiotics**<br>**n=36** | **P-value** |
| Microbiologically-confirmed infection | 24 (14.0) | 0 (0) | 0.02 |
| Respiratory cultures obtained | 66 (38.3) | 1 (2.7) | <0.01 |
| No growth | 53 [80.3] | 1 [100] | >0.99 |
| Positive | 13 [19.7] | 0 [0] | >0.99 |
| Blood cx obtained | 135 (78.5) | 14 (38.9) | <0.01 |
| No growth | 123 [91.1] | 14 [100] | 0.61 |
| Positive | 12 [8.9] | 0 [0] | 0.61 |
| Influenza PCR obtained | 75 (43.6) | 13 (36.1) | 0.46 |
| Strep pneumoniae urinary antigen obtained | 82 (47.7) | 6 (16.7) | <0.01 |
| Positive | 2 [2.4] | 0 [0] | >0.99 |
| Negative | 80 [97.6] | 6 [100] | >0.99 |
| Legionella urinary antigen obtained | 87 (50.5) | 6 (16.7) | <0.01 |
| MRSA nares obtained | 73 (42.4) | 2 (5.6) | <0.01 |
| Positive | 4 [5.5] | 0 [0] | >0.99 |
| Negative | 69 [94.5] | 2 [100] | >0.99 |

The most frequently prescribed initial antibiotics were ceftriaxone in 113 (66%) patients and azithromycin in 99 (57%) patients for community acquired pneumonia (CAP), followed by an anti-pseudomonal beta lactam (23%) and vancomycin (23%) for hospital acquired pneumonia (HAP) or ventilator associated (VAP) (Figures 1 and 2). The median time from SARS-CoV-2 collection to antibiotic initiation was one day (IQR 1-4.5 days). Median duration of therapy for the initial course of antibiotics was five days (IQR 2-6 days). In 41 (24%) patients, the initial regimen was changed; 17 patients were escalated to broader therapy (42%) and 15 patients were de-escalated to narrower therapy (37%). Nine (22%) had another regimen change such as intravenous to oral conversion or transition to another agent with similar spectrum of activity and not based on positive culture result.

**Figure 1:**
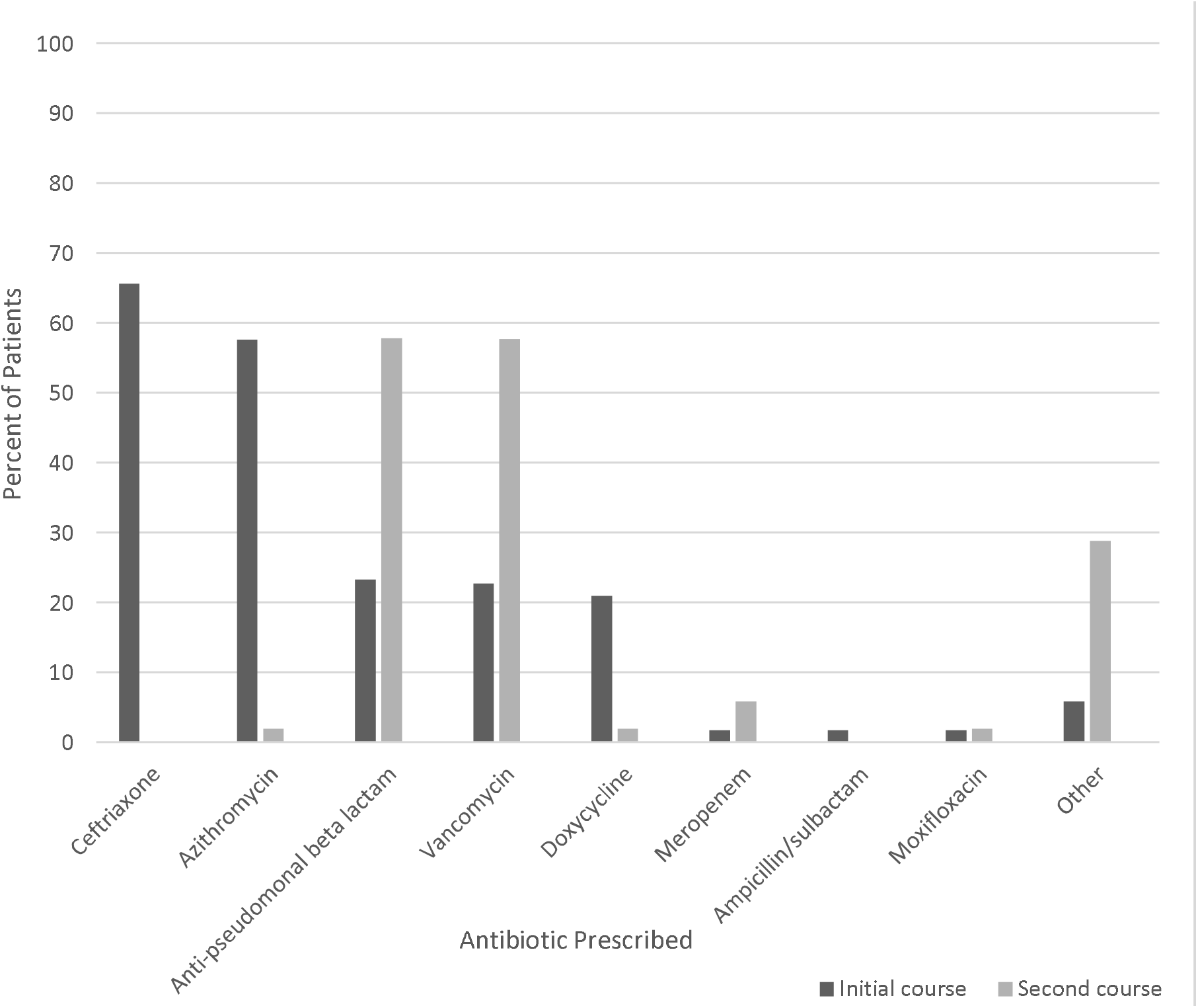
Antibiotic Prescribing Trends.

**Figure 2:**
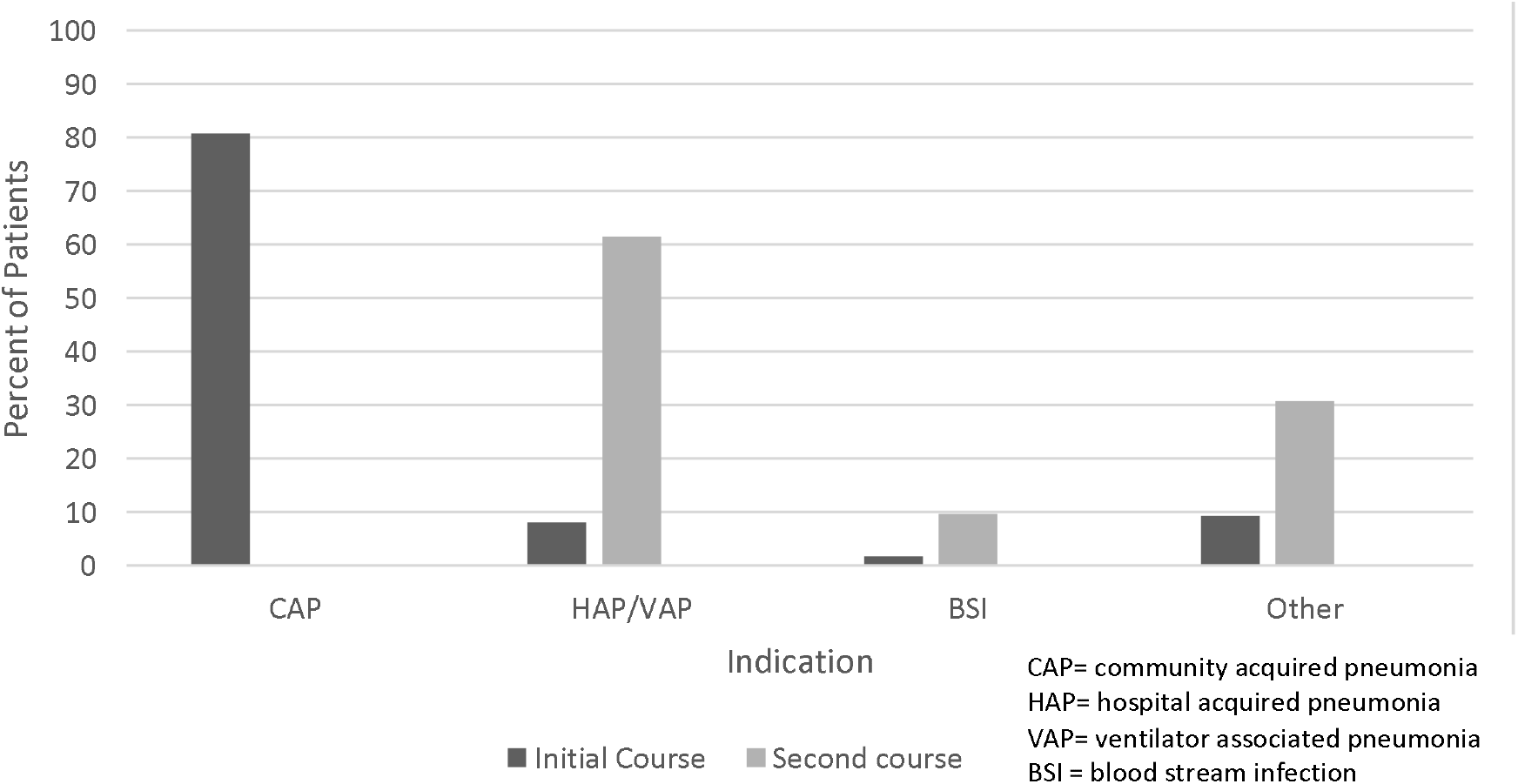
Indications for Antibiotic Prescribing.

Fifty-two patients (30%) received a second course of antibiotics and 19 patients (11%) received more than 2 antibiotic courses during hospitalization. The most frequently prescribed antibiotics included an anti-pseudomonal beta lactam (58%) and vancomycin (58%) for HAP/VAP (Figures 1 and 2). The median duration of second course antibiotics was 6 days (IQR 3-8.25). Duration of admission, ICU admission, ICU length of stay (LOS), mechanical ventilation, microbiologically-confirmed infection, and duration of initial course of antibiotics were all risk factors for receiving more than one course of antibiotics (Table 3). Of patients with microbiologically-confirmed infection who received additional antibiotic courses, 14 (70%) received treatment for HAP/VAP, 4 (20%) received treatment for bloodstream infection and 2 (10%) received treatment for other infections unrelated to cultures.

**Table 3:** Risk factors associated with receiving more than 1 course of antibiotics.

|  |  |  |  |  |  |  |
| --- | --- | --- | --- | --- | --- | --- |
| **Variable** | **Univariate Analysis** |  |  | **Multivariate analysis** |  |  |
|  | **1 antibiotic course**<br>**n= 120 (%)** | **> 1 antibiotic course**<br>**n=52 (%)** | **P value** | **Odds ratio** | **95% CI** | **P-value** |
| Length of stay, days (median, IQR) | 8 (4-15) | 25 (18.75-37) | <0.01 | 1.01 | 0.99-1.04 | 0.32 |
| ICU admission | 51 (42.5) | 49 (94.2) | <0.01 | 1.96 | 0.38-11.10 | 0.42 |
| ICU length of stay, days (median, IQR) | 5 (3-9) | 18 (12-26) | <0.01 | 1.14 | 1.06-1.26 | <0.01 |
| Mechanical ventilation | 25 (20.8) | 45 (86.5) | <0.01 | 1.83 | 0.46-7.61 | 0.39 |
| More than 1 COVID-19 related admission | 7 (5.8) | 1 (1.9) | 0.44 |  |  |  |
| Microbiologically-confirmed infection | 4 (3.3) | 20 (38.5) | <0.01 | 3.31 | 0.86-15.26 | 0.10 |
| Duration of initial course of antibiotics, days (median, IQR) | 4 (2-5.25) | 5 (4-7) | <0.01 |  |  |  |
| Time from SARS-CoV-2 collect date to initial antibiotic start, days (median, IQR) | 1 (0-5) | 2 (0-2) | 0.51 |  |  |  |
| 28-day mortality | 33 (27.5) | 14 (38.9) | >0.99 |  |  |  |

()= Percentages unless otherwise specified

## Discussion

In this multi-center study of patients hospitalized with COVID-19, we found an overall high rate of antibiotic prescribing, with 83% of patients receiving antibiotics. Of these patients, 30% received at least one additional course of antibiotics within the same hospitalization, increasing the overall antibiotic exposure. Despite the high rate of antibiotic prescribing, there were low rates of bacterial coinfection, with only 12% of patients having microbiologically-confirmed infection for initial antibiotic use. These findings are similar to the results of previous studies, suggesting the need for strategies to help clinicians judiciously prescribe antibiotics in patients with COVID-19.^1-5, 15^ Previous studies conducted in the United States have shown that concomitant community acquired infections in COVID-19 patients are low based on microbiological data within the first 48 hours of admission.^4, 5^ In our study, we found only slightly higher rates of microbiologically-confirmed infections than in other US studies, despite the fact that we reported all microbiologic data throughout hospitalization. The higher rate may be explained by bacterial infections acquired during hospitalization. Patients with microbiologically-confirmed infections were more likely to receive more than one course of antibiotics, be admitted to the ICU, and have a longer length of stay. Additional courses of antibiotics were often prescribed for empiric or targeted treatment of nosocomial infection.

Early in the pandemic, little was known regarding COVID-19 presentation, disease progression and rates of bacterial co-infection. Thus, guideline recommendations were evolving throughout the study period. Initial recommendations often encouraged antibiotic use. Within the University of Rochester Medical Center, our practices and institutional guidelines recommended initiating empiric antibiotics in the majority of COVID-19 patients. As information became more available, both national and institutional guidance changed, recommending judicious use of antibiotics. Subsequently, our antibiotic prescribing decreased. This was reflected in the temporal decline in antibiotic prescribing we observed from March to May.

There were several indicators that serious illness increased the risk of antibiotic prescribing. Initial antibiotic prescribing was more likely in patients with elevated PCT, longer length of stay, ICU admission, longer ICU length of stay, and administration of off-label or investigational therapy for COVID-19. Additionally, all patients with ARDS or requiring more invasive O2 support received at least 1 course of antibiotics. These findings are consistent with previous studies which found patients were more likely to receive antibiotics if they had severe disease upon presentation.^4, 5^ To our knowledge, this is the first study to report rates of additional antibiotic prescribing beyond initial empiric therapy. Patients with COVID-19 often have a prolonged hospitalization due to the underlying disease process. Our median LOS was significantly longer in patients who received at least one antibiotic course (12 days vs 5.5 days, p < 0.001), and prolonged hospitalization may also increase the potential for additional antibiotic exposure. Thus, it is important to understand antibiotic prescribing trends and associated risk factors beyond the first course. Our study found that nearly one-third of all patients received two or more courses of antibiotics. Variables associated with receiving two or more antibiotic courses included: longer length of stay, ICU admission, longer ICU length of stay, and mechanical ventilation. This is likely explained as these are all risk factors for hospital-acquired illness and antibiotics were commonly prescribed for nosocomial infection (e.g. HAP/VAP).

Procalcitonin has been suggested as a potentially useful biomarker to differentiate bacterial and viral infections and assist with clinical decision-making. However, the ATS/IDSA CAP guidelines strongly advise initiating antibiotics based off clinical judgement rather than PCT alone.^13^ In alignment with these guidelines, our institution does not recommend routinely using PCT to guide antibiotic initiation for CAP or sepsis. In COVID-19 patients specifically, there are conflicting data regarding baseline PCT in COVID-19 patients.^4, 16^ One two-center study found that patients with PCT available prior to antibiotic initiation were less likely to be prescribed antibiotics.^17^ At our sites, the patients prescribed antibiotics had significantly higher baseline PCT, however the median was only 0.28. In a subgroup analysis focusing on patients who had a PCT of > 0.5, 94% of patients (41/44) received antibiotics. Rates of microbiologically confirmed infection were similar between patients with a PCT > 0.5 and the entire cohort (12%). It is unclear at this time the utility of using PCT to determine if there is bacterial co-infection with COVID-19, and in our study it did not appear to be predictive of confirmed infection.

Previous studies have shown high rates of broad-spectrum antibiotics for initial empiric therapy in COVID-19 patients.^1, 3^-^6, 8, 15^ However, in our patient population, most patients were treated initially for CAP. The most common agents included ceftriaxone plus azithromycin, which is consistent with the 2019 ATS/IDSA recommendations for CAP.^13^ It is worth noting that our institution did not routinely recommend use of azithromycin off-label for anti-inflammatory effects in COVID-19. During the initial antibiotic course, 37% of patients were de-escalated to narrower therapy. Not surprisingly, patients who received a second course of antibiotics were more likely to receive broad-spectrum antimicrobials; over half of patients who received more than one course of antibiotics were treated for HAP/VAP and prescribed an anti-pseudomonal beta lactam. Respiratory cultures were obtained in only 30.2% of patients, with a large number of patients receiving empiric treatment for CAP, HAP or VAP. In part, the low number of respiratory cultures obtained may be due to the symptomatology of disease, with the majority of patients experiencing dry, non-productive cough.^1^ Additionally, limited PPE early in the pandemic created additional challenges in obtaining respiratory cultures due to concerns of transmission and aerosolization. Also, the 2019 ATS/IDSA CAP guidelines do not recommend routine sputum gram stain and culture in patients with non-severe CAP or those without multi-drug resistant organism risk factors due to overall poor yield and detection of organisms.^13^ However, respiratory cultures are recommended in patients being treated empirically for MDR organisms, and in those being treated for HAP/VAP.^13, 14^ Respiratory cultures were not obtained in all patients who met these criteria. Obtaining respiratory cultures and other microbiologic data, when possible, on patients treated with broad spectrum antibiotics can potentially help with de-escalation of therapy.

One limitation of this study is its retrospective design. Laboratory and microbiologic data collected were not consistent amongst all patients. We found duration of admission and ICU length of stay to be associated with higher antibiotic use, however it is difficult to say that these are true risk factors for antibiotic prescribing. Instead, this difference was likely due to antibiotic prescribing being higher in patients with more serious illness, and perhaps longer admission leading to increased risk of nosocomial infections and subsequent antibiotic courses. Additionally, we did not perform a time adjusted analysis. Another limitation is that the vast majority of patients within our study received antibiotics which could have affected our ability to detect differences between the two groups. Early in the pandemic, most COVID-19 patients within our institution received antibiotics. While we saw a temporal decrease over time, it is difficult to assess risk factors for antibiotic use in our patients treated for COVID-19 early in the study when antibiotic use was routine.

While there were limitations of this study, there were also several strengths. This study was multi-site, including three hospitals within the University of Rochester Medical Center: two community hospitals and a tertiary academic medical center. Therefore, the results of this study may be generalizable to similar sites. Finally, as previously mentioned, this is the first study to assess overall antibiotic use throughout hospitalization, and determine risk factors for additional antibiotic courses. Many patients with COVID-19 have prolonged hospitalization, increasing the risk for antibiotic exposure. Our study examined antibiotic use that is more representative of hospitalized COVID-19 patients. Based on the results of this study, our institution plans to examine how other factors may increase or decrease the risk of antibiotic prescribing, as well as trend our use of antibiotics throughout the pandemic.

## Conclusions

Our study found high rates of antibiotic prescribing, despite low rates of respiratory cultures and confirmed microbiologic infections. Nearly one-third of patients were treated with more than one antibiotic course during hospitalization. These data highlight a role for antimicrobial stewardship during the COVID-19 pandemic. Further studies are needed to examine the impact of antimicrobial stewardship initiatives in patients with COVID-19 and assess if antibiotic prescribing practices have changed with increasing information related to bacterial co-infection that wasn’t available at the beginning of the pandemic.

## Data Availability

This research is based on data collected within our institution. No databases were used for the completion of this project.

## Acknowledgements

The authors thank Raquel Roberts, PharmD for her assistance with data collection. There was no funding for the study.

